# Performance of a Semi-Automated Hierarchical Rest Interval Detection Pipeline (actiSleep) for Wrist Actigraphy in Adolescents

**DOI:** 10.64898/2026.03.05.26347744

**Authors:** Adriane M. Soehner, Nicholas Kissel, Brant P. Hasler, Peter L. Franzen, Jessica C. Levenson, Duncan B. Clark, Daniel J. Buysse, Meredith L. Wallace

## Abstract

Actigraphy is a popular behavioral sleep assessment tool in research and clinical practice. Hierarchical hand-scoring approaches remain the standard for actigraphy rest interval estimation, but can be impractical for large cohort studies and suffer from reproducibility problems. We developed a semi-automated pipeline (actiSleep) to set rest intervals consistent with best-practice hand-scoring algorithms incorporating event marker, diary, light, and activity data. To evaluate actiSleep performance, we used data from an observational study of 51 adolescents (14-19yr), with and without family history of bipolar disorder. Participants completed 2 weeks of wrist actigraphy and daily sleep diary. We first hand-scored records using a standardized hierarchical algorithm incorporating event marker, diary, light, and activity data. We then compared the hand-scored rest intervals to those from actiSleep and two automated activity-based algorithms (‘Activity-Merged’, ‘Activity-Only’). Activity-Only used activity-based sleep estimation and Activity-Merged joined closely adjacent rest intervals. For rest onset, rest offset, and rest duration, all algorithms had strong mean agreement with hand-scoring: actiSleep estimates were within 1-3 minutes, Activity-Merged within 2-4 minutes, and Activity-Only within 7-14 minutes. However, actiSleep had notably better (narrower) margins of agreement with hand-scoring, as evidenced by Bland-Altman plots, and greater positive predictive value and true positive rates for rest detection, especially in the 60 minutes surrounding the onset and offset of the rest interval. The actiSleep algorithm successfully estimates actigraphy rest intervals comparable to hand-scoring while avoiding pitfalls of activity-only algorithms. actiSleep has potential to replace hand-scoring for research in adolescents but requires further testing and validation in other samples.

## INTRODUCTION

Due to its convenience, accessibility and affordability, wrist actigraphy has emerged as a widely used behavioral sleep estimation approach in both research and clinical settings. While polysomnography remains the gold standard objective assessment of physiological sleep, the expense and expertise required for its implementation as well as its obtrusiveness make it an impractical approach for long-term monitoring of naturalistic sleep patterns. As a result, wrist actigraphy has been increasingly used in large cohort studies of sleep and health. However, automated actigraphy sleep scoring has known biases, and the alternative – manual scoring – remains burdensome and inefficient. New approaches that blend the ease of automated scoring with the precision of manual hand scoring are needed to ensure actigraphy monitoring is both an accurate and feasible behavioral sleep estimation approach.

Wrist actigraphy uses an accelerometer embedded in a watch-like device to continuously measure wrist motion (“activity”). Research-grade actigraphy devices can record activity for weeks to months on a single charge. After initial signal processing that generates epoch-level time series (e.g., activity counts within bins of 30- to 60 seconds), automated sleep estimation algorithms (e.g., Cole-Kripke^1^, Sadeh^2^, HDCZA^3^, Oakley^4^) can be applied to classify each epoch as either sleep or wake. Subsequently, ‘rest intervals’ (spans of time when individuals are *trying* to sleep) as well as specific ‘sleep intervals’ (spans of time when individuals are *actually* sleeping) can be automatically set. While efficient, this automated approach to setting the rest interval can result in rest intervals being misclassified or overestimated. Moreover, immobile wakefulness is prone to misclassification as sleep^5^, particularly when applied across the 24-hour day (i.e. without the confines of a valid rest interval) and/or among individuals with poor sleep^6–8^.

To address these issues, published best practices^9^ recommend that supplemental information from the sleep diary or sensors embedded in the actigraphy device itself (light sensor, event marker) are considered in combination with activity-based sleep estimation to manually identify the rest interval with greater confidence. Sleep is then estimated within this fixed rest interval using the automated activity-based algorithm.

Supplemental indicators for rest interval determination are especially important for improving the validity of sleep latency and efficiency estimates in home-based actigraphy studies^10^ because computing these metrics require the precise time that an individual begins to try to fall asleep (i.e., the start of the rest interval).

However, in practice, there is considerable variation in how multiple indicators of rest are weighed in actigraphy scoring, likely contributing to reduced reproducibility.

Standardized approaches integrating supplemental information into actigraphy hand-scoring have been recommended for rest interval detection^11–14^ and have great potential to improve reproducibility of actigraphy-based sleep studies. One popular standardized algorithm that showed promising inter-rater reliability was proposed by Patel et al.^11^. This scoring algorithm leverages a rigorous combination of hierarchical rules for weighing the importance of each ‘indicator’ of rest (marker press, sleep diary, light intensity, activity), as well as the degree of concordance between these indicators, to determine the onset and offset of each rest interval.

However, hand-scoring using current standardized approaches can be time-intensive and impractical in large cohorts.

As another alternative to inefficient hand-scoring, several sophisticated open-source pipelines (e.g., GGIR^15^, pyActigraphy^16^) and machine-learning methods^17–20^ have been developed to automate actigraphy scoring at a large scale. These systems primarily offer the options for activity-based sleep estimation only or use a manually entered ‘guider’ (typically, the sleep diary) to set the rest interval. None of these methods, to our knowledge, incorporate *multiple* different data sources used in standardized hand-scoring (e.g., marker press, sleep diary, light intensity), which may limit their accuracy, sensitivity, specificity for detecting rest intervals in field-based studies. An automated rest interval detection algorithm for actigraphy that incorporates multiple data sources (activity, light, marker press, sleep diary) and mimics hand-scoring decision rules could drastically reduce effort required to rigorously define rest intervals and enhance reproducibility.

We developed a semi-automated algorithm called actiSleep that mimicshierarchical decision rules taught to human scorers, and which incorporates diary, light, marker presses, and activity. Using a convenience sample of adolescents at high and low risk for bipolar disorder with a range of mental health and sleep problems, we evaluated the performance of actiSleep relative to hand-scoring. For comparison, we also evaluated a fully automated version of actiSleep requiring only activity data that mimics the way hand-scorers merge closely adjacent rest intervals (‘Activity-Merged’) and the Phillips-Respironics automated activity-based sleep algorithm (‘Activity-Only’). Because actiSleep is designed to improve accurate and reliable identification of the rest interval and incorporates the same information available to hand-scorers, we posited that it would out-perform the Activity-Merged and Activity-Only algorithms for estimating Rest Onset, Rest Offset, and Rest Duration.

## METHODS

### Participants

Our sample comprised adolescents recruited for a sleep and neuroimaging pilot study, and included both typically developing healthy controls (Low-Risk; n=29) and teens with elevated risk for bipolar disorder based on parental history of the diagnosis (High-Risk; n=22). <u>All participants</u> were: a) 14.0-19.11 years old; b) enrolled during the high school/college academic year; c) right-handed; d) MRI eligible (i.e., no ferromagnetic metal in the body); e) free of major systemic medical illnesses, neurological disorders, history of head trauma, and intellectual disabilities (parent report); f) free of organic sleep disorders (e.g., apnea, narcolepsy, restless legs syndrome, REM behavior disorder) per a locally-developed semi-structured interview; and g) not pregnant or breastfeeding (self-report and/or urine test).

<u>Low-Risk participants</u> and their parents were free of psychopathology; adolescents a) had no current or lifetime history of psychiatric conditions based on the Kiddie Schedule for Affective Disorders and Schizophrenia (KSADS^21^); b) had no first degree family history (parent, sibling) of a psychiatric condition based on the Structured Clinical Interview for the DSM-5, Research Version (SCID;^22^) and Family History Interview (FHI^23^); and c) were not taking psychotropic or hypnotic medications. <u>High-Risk participants</u> could have non-severe psychiatric conditions and had a parent with bipolar disorder; they were: a) free of lifetime bipolar disorder, lifetime psychotic disorder, and alcohol/substance use disorders in the past 3 months (KSADS^21^); b) had at least one parent diagnosed with DSM5 type 1, type 2, or Other Specified bipolar disorder (SCID^22^ and/or FHI^23^); and c) could be taking psychotropic medications if the medication type and dosage were stable for at least 2 months.

Participants were recruited via community advertisement, local clinics, and a participant registry. The University of Pittsburgh Human Research Protection Office approved the study; all participants provided written informed consent or assent. All participants with at least 1 week of wrist actigraphy data were included in the present analysis.

### Psychiatric Screening Measures

DSM-5 psychiatric disorders were assessed using the *Kiddie Schedule for Affective Disorders and Schizophrenia* (KSADS)^21^ in offspring participants, the *Structured Clinical Interview for DSM-5* (SCID)^22^ in the parent attending the study visit, and the *Family History Interview* (FHI)^23^ in first-degrees relatives of the offspring participant. The *Pittsburgh Sleep Diagnostic Interview* (PSDI) assessed for offspring sleep disorders using DSM-5 criteria; this is a locally developed sleep disorders interview.

### Actigraphy

Participants were asked to wear a wrist accelerometer (Actiwatch Spectrum; Philips Respironics, Bend, OR) continuously on the non-dominant wrist for 14 days. The Actiwatch Spectrum includes an accelerometer to assess activity and a tricolor light sensor. The light sensor captures blue (400-500 nm), green (500-600 nm), and red (600-700 nm) which are integrated to capture illuminance of environmental white light (range 0.1-200,000 lux). Light and accelerometer data were collected in 60-sec epochs. Participants were instructed to wear their Actiwatch continuously except in salt water and during activities that may damage the device (e.g., contact sports). For each rest interval, participants were asked to indicate ‘good night time’ (i.e., the beginning of a sleep attempt) and ‘get up time’ (i.e., physically getting up from a sleep attempt) via button presses on the Actiwatch.

### Sleep Diary

Daily self-report sleep diary captured standard data elements recommended by the consensus sleep diary^24^ (e.g., bedtime, lights off time, sleep latency, wake after sleep onset, number of nocturnal awakenings, sleep offset time, get up time) for each bout of sleep. In addition, participants were asked to classify each sleep bout as a nap or their main sleep interval for the day. Links to the sleep diary were distributed to participants via text message each morning at their preferred time, based on habitual weekday and weekend rise times.

Sleep diaries had a 48-hour response window (i.e., participants could only report on their sleep patterns for that day and the prior day).

### Study Design

Adolescent participants (and a parent, if <18yr) completed consent/assent and an evaluation to determine study eligibility that included psychiatric diagnostic interviews (SCID, KSADS, FHI), a sleep diagnostic interview, and questionnaires. Eligible participants were invited to complete 2 weeks of continuous actigraphy monitoring and an online sleep diary each morning. Sleep monitoring occurred during the school year only.

### Scoring Algorithm Components

#### Sleep Estimation Algorithm

In Actiware software version 6.0.9, activity-based sleep estimation was performed using the Oakley medium wake threshold setting for the main rest interval (10 minutes immobile time for sleep onset and offset). For detection of minor rest intervals (naps), we used a medium wake threshold with a 45-minute minimum, based on a prior validation study^25^. Off-wrist and invalid time were automatically converted to Excluded intervals. Actigraphy ‘tracking days’ were defined as noon-to-noon.

#### Rest Indicator Definitions

**Table 1** summarizes the rules used to establish the clock time of rest onset and offset for each of the four rest interval indicators (activity, light, sleep diary, and marker press). These rules were used for both hand-scoring and automated scoring.

**Table 1.** Actigraphy Indicator Definitions.

| Indicator | Rest Onset Definition | Rest Offset Definition | Indicator Validity Rules |
| --- | --- | --- | --- |
| Marker Press | Marker press near a notable drop in activity, light, or Sleep diary Good Night time | Marker press near a notable rise in activity, light, or Sleep Log Get Up time | • If there are multiple marker presses in close succession, use the latest<br>• Errant marker presses can be disregarded |
| Sleep diary | Self-report Good Night time | Self-report Get Up time | • Disregard if sleep diary completed > 2 days after tracking day |
| Light | Sudden, large drop in white light intensity, falling to <1 lux on average for >30min | Sudden, large increase in white light intensity that rises to over 1 lux on average for >30min. If 1 lux criteria was not met, we used a personalized threshold<sup>a</sup>. | • Gradual increase white light intensity in the early morning was considered 'sunrise' |
| Activity | Actiware medium-threshold (10 immobile minutes) rest interval start | Actiware medium-threshold (10 immobile minutes) rest interval end | • Disregard if there was indication of an early, brief wake-up (i.e., a trip to the restroom) before the final get-up time. |
| <sup>a</sup>Personalized thresholds were used only for actiSleep. Details provided in the actiSleep algorithm section. |  |  |  |

#### Indicator Hierarchy Rules

This hierarchy is used in hand-scoring and actiSleep to determine how to prioritize and integrate information from each of the four indicators when determining rest onset and offset (**Table 1**).

*Rest Onset (Good Night Time).* The indicator hierarchy was Marker > Sleep Diary > Light > Activity, informed by the Patel algorithm^11^. The beginning of the rest interval was defined as the start of the sleep attempt (“lights off”, “good night time”). ‘Subjective’ indicators were prioritized (marker press, sleep diary) due to the subjective nature of attempting sleep. We counted the number of pairs indicators that were ‘concordant’, as defined as occurring within 15-minutes of each other. If at least 2 indicators were concordant, a preliminary ‘winner’ indicator was selected based on the hierarchy. If 2 pairs of indicators were concordant (e.g., both marker and light suggested time in bed was at 22:00 while both diary and activity both suggested 23:30), the highest ranked indicator (i.e., marker) was the preliminary ‘winner’ indicator. If no indicators were within 15 minutes of one another, concordance was re-evaluated using a 30-minute window. If no indicators overlapped within 30min, the best available indicator in the hierarchy was selected as the preliminary ‘winner’ indicator.

After a preliminary ‘winner’ indicator was determined, two special cases were then assessed. If a marker press occurred up to 120min after the preliminary ‘winner’ indicator, Rest Onset was shifted later to the marker press time (Case 1). The rationale was that the marker press is likely only to occur if the participant is not yet asleep. Next, if the light indicator occurred up to 120min after the preliminary ‘winner’ indicator (including marker press in Case 1), Rest Onset was shifted to the light indicator (Case 2). The rationale is that the sleep attempt has unlikely to have begun if the light has not been turned off. These special cases were applied to our cohort on participants accounts of what patterns best captured their Good Night Time; actograms were reviewed on one-on-one with researchers and participants at the conclusion of actigraphy monitoring. Special cases can be readily modified in the actiSleep pipeline; special cases may need to be developed for each population or removed for greater generalizability across populations.

*Rest Offset (Get Up Time).* The indicator hierarchy was Activity > Light > Marker > Sleep Diary, akin to the Chow algorithm^13^. Because we were interested in Get Up time as the end of the rest interval, objective indicators were prioritized (activity, light) to better capture the behavioral act of rising from a recumbent resting position. Light was omitted if a gradual rise in light was detected, reflecting ‘sunrise’. Similar to Rest Onset, concordance between indicators was assessed at 15-minute and 30-minute intervals for Rest Offset. If at least 2 indicators were within 15 minutes of each other, a preliminary ‘winner’ indicator was selected based on the hierarchy. If no indicators were within 15 minutes of one another, a 30-minute comparison window was used. If 2 pairs of indicators were concordant (e.g., both marker and light suggested get up time was at 7:00 while diary and activity both suggested 6:10), the highest ranked indicator (i.e., activity) was the ‘winner’ indicator. If no indicators overlapped within 30min, the best available indicator in the hierarchy was selected as the preliminary ‘winner’ indicator.

#### Rest Interval Merging

Rest intervals were merged under two sets of conditions. The first merging approach combined rest intervals that were closely adjacent. Rest intervals were merged if they occurred within ≤1 hour of one another (i.e., rest offset of interval 1 was within 1 hour of rest onset of interval 2. The second merging approach aimed to capture lengthier nocturnal awakenings that bifurcated the presumed ‘main’ rest interval. The median main rest interval onset and offset time across the tracking period were estimated based on the Oakley algorithm described in the section above. If a period of extended wakefulness (i.e., one that was identified as an “active interval” by automated scoring) occurred between the median rest onset and median rest offset time, then the surrounding rest intervals were merged.

#### Rest Interval Deletion

If a minor rest interval (nap) had <15min of detected sleep, the interval was deleted, unless the interval was corroborated by marker press and/or sleep diary. This rule was put in place to minimize the inclusion of inactive wakefulness as spurious naps.

#### Main Sleep and Nap Interval Definitions

The ‘main’ sleep interval was preliminarily defined as the longest rest interval of the day. Then, the sleep midpoint for each day during the tracking period was calculated and the median sleep midpoint of the tracking period was identified. The ‘main’ sleep interval for each day was then updated to correspond to the rest interval whose midpoint was closest to the calculated median midpoint. This process accounted for situations where the sleep interval occurring at the typical main rest interval time for a participant (e.g., at night) might be shorter than a daytime nap.

#### Exclusion of Invalid Days

Days with either (i) more than 240 minutes of invalid time or (ii) more than 15 minutes of invalid sleep based on prior work^11^ were deemed invalid and excluded from all analyses.

### Scoring Algorithms

Figure 1 provides an overview of the four different scoring algorithms that were compared. Hand-scoring was performed by an expert scorer (AMS) prior to comparison with the other three algorithms. It was used as reference scoring. actiSleep is our primary algorithm of interest that we are testing relative to hand scoring. We also considered two fully automated comparator algorithms: Activity-Merged and Activity-Only. Both hand-scoring and the actiSleep algorithm are based on a common set of actigraphy scoring rules; minor differences and the corresponding rationale are summarized in the actiSleep workflow.

**Figure 1.**
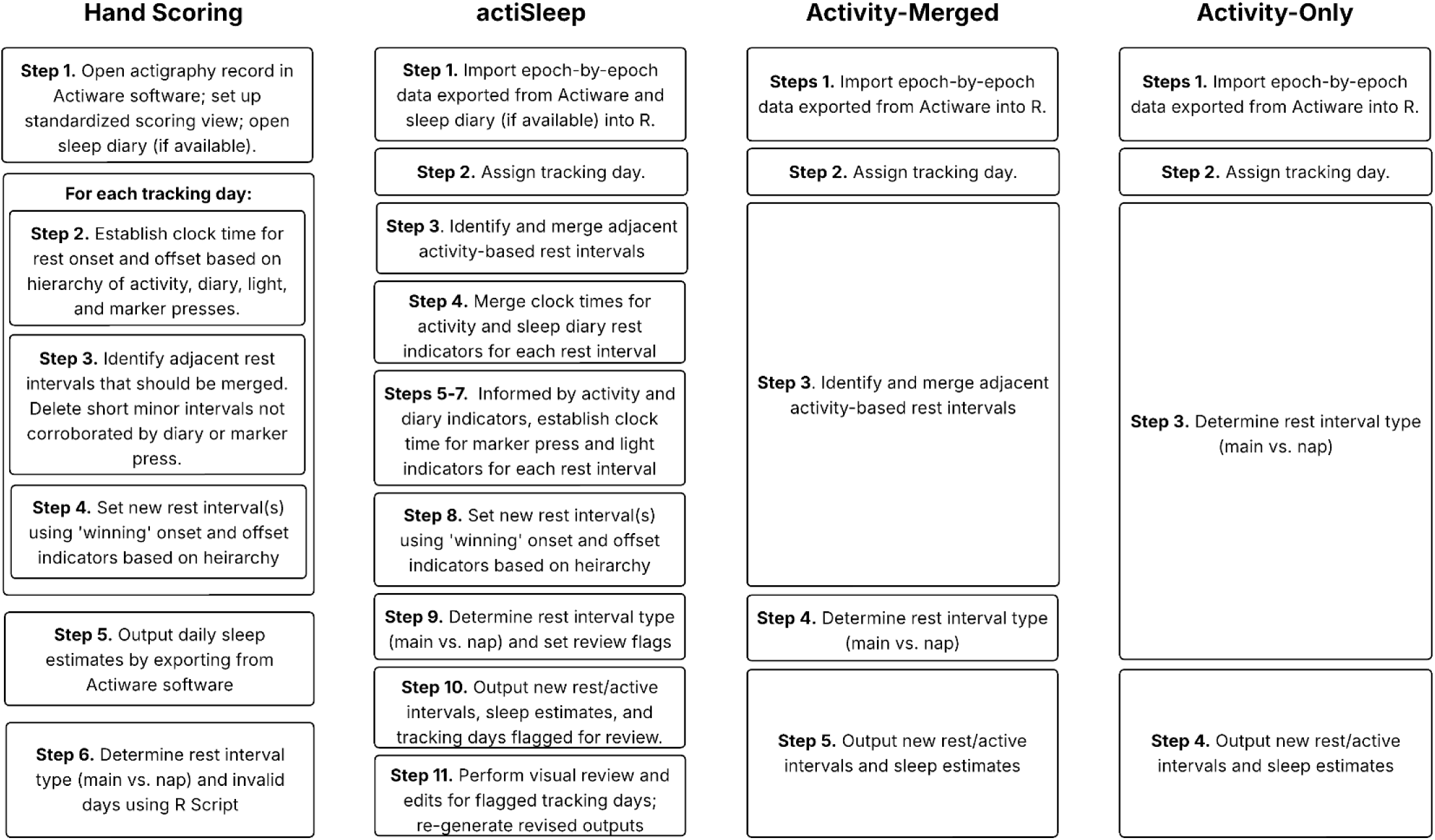
High-level overview of the steps involved in each of the four scoring appraoches compared

### Hand-scoring

Hand-scoring uses activity-based rest intervals, light, diary and marker data to define rest intervals, and was used as the reference algorithm. The scorer first initializes the record in Actiware and sets the actogram view to seven noon-to-noon tracking days with activity time series, white light time series, and the activity-determined rest intervals visible, then opens the participant’s sleep diary spreadsheet (Step 1). The next steps (2-4) are performed for each tracking day for each rest interval. Clock times are determined for rest onset and offset based on activity-based sleep estimation, sleep diary, marker press, and light intensity, as in **Table 1**, then the ‘winner’ indicator is determined based on the hierarchy rules noted above (Step 2). The scorer assesses whether any rest intervals should be merged or deleted, based on the rules noted above (Step 3).

Rest-interval merging/deletion occurs in parallel with considering indicator concordance during hand-scoring, as rest intervals without corresponding marker press, diary, or light indicators guide the scorer to the ‘true’ rest interval. The algorithm-determined rest interval onset and offset are marked in the actogram, and a new rest interval is populated (Step 4). Daily sleep estimates are exported from Actiware (Step 5), then interval type (main, nap) and invalid days (Step 6) are defined in a custom R script based on the rules noted above.

### actiSleep

<u>actiSleep</u> is a semi-automated algorithm that uses activity, light, diary and marker data that closely mimics the process of hand-scoring. After importing data and assigning tracking days (Steps 1-2), activity-based rest intervals as defined by Actiware are merged (Step 3). For each rest interval, clock times are merged across activity, diary, maker, and light (Step 4). Rest interval onset and offset times for marker presses and light are then iteratively determined (Steps 5-7). Comparing across all available indicators for each day, the clock time for rest onset and offset is determined based on a prescribed hierarchy (Step 8; see **Table 1**) and interval type is determined (Step 9). Finally, we set flags for review (Step 10) and output the new rest and active intervals, along with sleep estimates (Step 11). This semi-automated actiSleep algorithm adhered to all rules described above, plus ‘flagging’ of tracking days requiring visual review and hand-scoring, described in the section below. The actiSleep algorithm is implemented in R and is available on https://github.com/nkissel/actiSleep.

To run actiSleep, only an actigraphy epoch-by-epoch csv data file is required in addition to the actiSleep R function. For this implementation, epoch-by-epoch csv files were exported from Actiware, containing the epoch-level data (timestamp, activity, light, marker presses, interval type, nonwear), rest-interval summary statistics, and device properties. Sleep diary input is optional. If available, the sleep diary file should be in csv format and contain ID and good night time and get up time date-time stamps for each rest interval, for all participants. Details on each specific step follow.

*1. Import data*: actiSleep reads in the actigraphy and (if available) sleep diary data files provided by the user.
*2. Assign ‘tracking day’:* Using an anchor date (the date defined as the official ‘day one’ for the participant), a common tracking day number is assigned to each row of data in both the actigraphy and sleep diary files. ‘Tracking days’ are defined as noon-to-noon intervals. The tracking day number (according to the anchor date) is assigned to the *beginning* of each rest interval. Invalid days are excluded from further analysis. Any days of actigraphy or diary occurring prior to tracking day 1 are excluded from further analysis.
*3. Identify and merge activity-based rest intervals (actigraphy file only):* Rest intervals are first determined and processed based on activity-based sleep estimation. Rest interval merging rules are applied, like hand-scoring. However, this step occurs earlier than hand-scoring and merging rules are applied recursively, otherwise sleep diary and marker press indicators will not appropriately align in Steps 4-5. A list of updated rest intervals for activity-based rest onset and offset date-times is generated.
*4. Combine activity and sleep diary-based rest interval times:* Activity and sleep diary rest interval onset and offset date-times are merged into a single dataset, by ID and tracking day.
*5. Identify rest intervals based on marker presses:* We identify marker presses occurring within 60 minutes of either activity or diary rest onset or offset. As there can be extra marker presses, constraining the search space helps identify the marker press most likely to be concordant with other indicators. If there are still multiple marker presses within this interval, the latest in the series is selected for rest onset, and the earliest in the series for rest offset. Identified marker press rest interval onset and offset times are merged with activity and sleep diary rest interval times.
*6. Identify rest intervals based on light:* Light indicators for rest onset/offset are determined using the criteria in **Table 1** plus a personalized threshold to account for individual differences in light exposure patterns, which may stem from differences in the sleep environment or time of year. For a given day the personalized threshold was defined using three steps. (1) Calculate the smoothed log-light for each epoch using lowess. (2) Identify the percentile of the smoothed log light corresponding to the percentage of activity-based sleep for that day. Record the corresponding lux value. (3) Identify the time of day that the light crosses the lux threshold and it stays above (for activity-based rest onset) or below (for activity-based rest offset) it for 30 minutes.
*7. Update marker press rest intervals:* Marker press rest interval times are updated, now including light onset/offset in the search window.
*8. Select ‘winner’ indicator for each rest interval:* Based on the concordance algorithm described above, the ‘winner’ indicator is selected for each rest interval onset/offset.
*9. Determine interval type and set flags:* Each rest interval is marked as a nap or main interval, based on the definition above. A subset of tracking days is flagged for manual review based on main rest interval characteristics. Criteria were based on the age range of our sample and likely require adjustment based on the sample under study. Flags included: **short main interval duration** (rest interval that is < 3 hours long and a maximum indicator discrepancy > 1 hour), **long main interval duration** (rest interval > 14 hours and a maximum indicator discrepancy > one third of the interval duration), **long sleep latency** (rest interval has sleep latency ≥ 180 minutes), **missing sleep intervals** (tracking day has no sleep intervals), and **close adjacent rest intervals** (rest intervals that are within 2 hours of one another, where the wakeful period occurs between 10PM and 6AM).
*10. Generate outputs:* For each ID, we generate a long-form data set showing each rest and active interval across tracking days, plus the onset and offset times defining the rest and active intervals. For rest intervals, we show its determination of main vs. nap, plus summary statistics including sleep onset and offset times, sleep latency, wake after sleep onset, and sleep duration. We also identify whether a particular rest interval was flagged for review, the reason for flagging, and indicate any invalid/excluded days.
*11. Visual review of flagged tracking days.* A trained reviewer performed a visual review of actograms with flagged tracking days to determine if manual edits were required (see Figure 2). Our data set included 575 non-excluded tracking days and 29 flagged days. A total of 19 flagged days had rest intervals were hand-scored by the visual reviewer (AMS); the other 10 were determined to be correct. New rest onset and offset times are recorded and summary statistics are recalculated. Final data are re-generated and saved.

**Figure 2.**
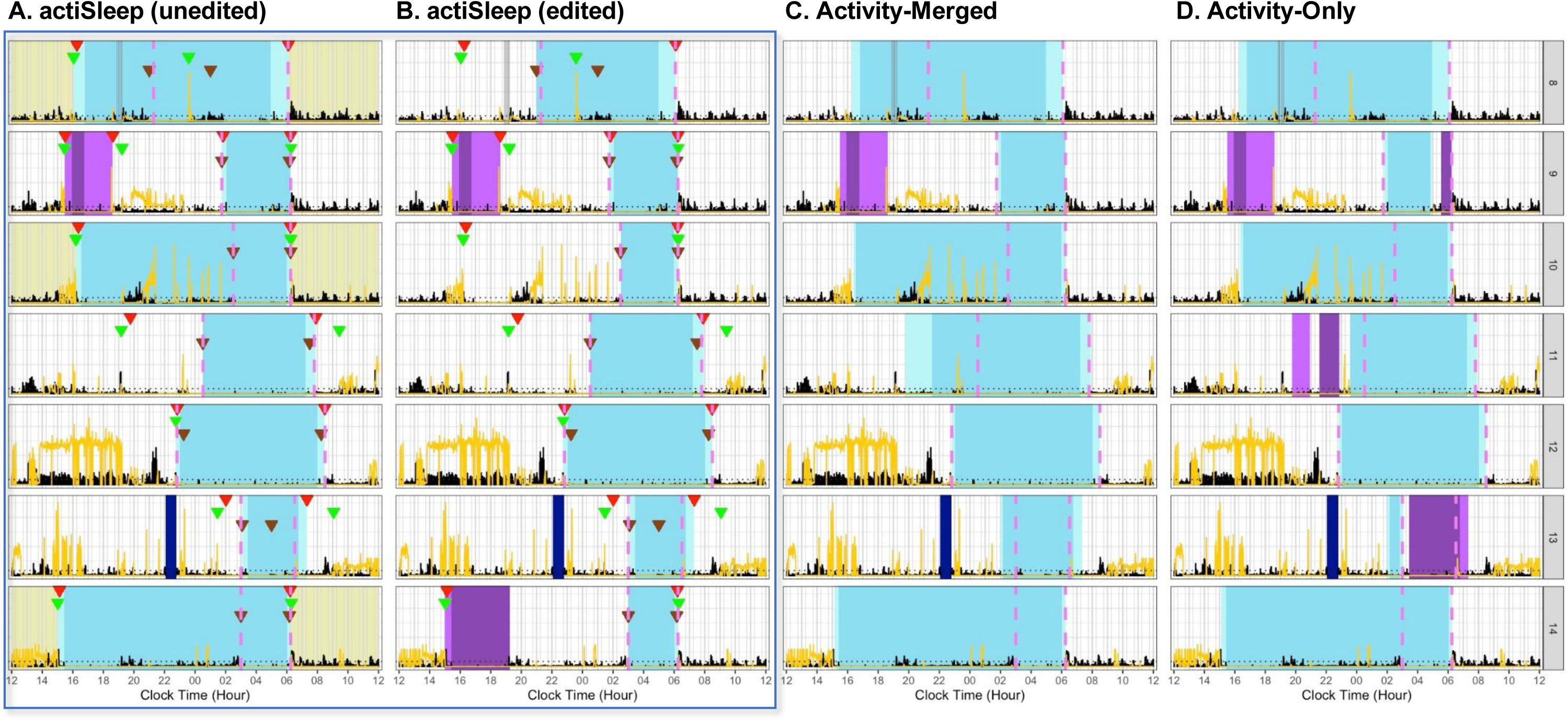
Example actograms generated from actiSleep (before [**A**] and after editing [**B**]), Activity-Merged [**C**], and Activity-Only [**D**] for a single subject. Each row reflects a noon-to-noon tracking day. Light yellow shading of a tracking day reflects a day flagged for manual review. Activity (black) and light (yellow) are plotted by epoch. Each tracking day, light blue shading reflects a rest interval; the mid-blue shading within the rest interval reflects the sleep interval; purple shading reflects a nap. Rest interval indicators are plotted at the start and end of each rest interval (dark blue = marker press, brown = sleep diary, green = light, red = activity). The pink dashed line indicates hand-scoring rest interval start and end. Navy blue shading reflects a non-wear interval.

### Activity-Only

Activity-Only is the automated sleep estimation algorithm, described above, implemented in Actiware.

This algorithm was included to provide a baseline for the accuracy of a fully automated actigraphy scoring algorithm. After importing data and assigning tracking days (Step 1), we determined the interval type (Step 2) and then output rest and active intervals and sleep estimates (Step 3).

### Activity-Merged

Activity-Merged is a fully automated algorithm that uses only activity data, but which includes some functionality of actiSleep by merging rest intervals that are close together. This algorithm was included to evaluate the extent to which the incorporation of diary and marker press further added value to rest interval detected within the actiSleep algorithm. After importing the data and assigning tracking days (Step 1), we use the same process as in actiSleep to merge activity-based rest intervals (Step 2). Interval types are then determined (Step 3) and we output new rest and active intervals, along with sleep estimates (Step 4).

### Analyses

In preliminary analyses, we summarized sample characteristics and the number of rest onset and offset periods with observed diary, light, activity and marker push indicators. We also summarized the number and types of flags identified in the sample, and the number of these flags that resulted in altering the algorithm rest onset of offset determination.

### Algorithm Agreement

Bland-Altman plots were used to visualize the level agreement between each rest interval detection algorithm (Activity-Only, Activity-Merged, actiSleep) relative to hand-scoring for the main rest interval. Primary outcomes included rest interval onset, rest interval offset, and rest interval duration. Secondary outcomes were: sleep onset, sleep offset, sleep latency, wake after sleep onset, snooze time, total sleep time, and sleep efficiency. The mean difference (± 1.96 standard deviation of the mean difference) between each algorithm and hand-scored measures were shown on the y-axis and plotted against the hand-score measures on the x-axis. In secondary analyses, we examined and compared algorithm agreement for high-risk versus low-risk groups by color coding the Bland Altman plots accordingly (see **Supplement**).

Paired t-tests or Wilcoxon signed-rank tests were used to evaluate differences in sleep outcomes between each algorithm and hand-scoring for the main sleep interval; main analyses focused on primary and secondary sleep outcomes averaged across the full tracking interval. For primary and secondary outcome, α was set to 0.05, and Benjamini-Hochberg multiple comparison correction was applied across all tests. P-values <0.05 after multiple comparison correction were considered significant.

### Epoch-level Accuracy

For a given scoring approach (hand-scoring, actiSleep, Activity-Merged, Activity-Only), all epochs between the rest onset and offset times are classified as rest (1) and all other epochs are classified as active (0). We were interested in the ability of our algorithms to identify the rest interval, not to evaluate the performance of the Oakley sleep estimation algorithm used by Phillips Respironics. We evaluated the performance of the three semi/fully automated algorithms’ classification of active and rest epochs relative to hand-scoring classifications, which were taken as the ground truth. Because of the relatively large number of true active epochs relative to true rest epochs, we focused on the positive predictive value (PPV) and true positive rate (TPR). PPV summarizes the proportion of the algorithm-classified rest intervals that are true hand-scored-classified rest intervals, quantified as 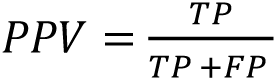 where TP is the number of ‘true’ rest intervals (defined by hand-scoring) correctly identified as rest by the algorithm and FP is the number of ‘true’ active intervals (defined by hand-scoring) incorrectly identified as rest by the algorithm. In contrast, TPR addresses the sensitivity or ‘recall’ of rest epoch classification, quantified as 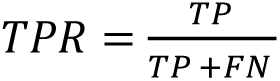 where TP is defined as before and FN is the number of ‘true’ rest intervals (defined by hand-scoring) incorrectly labeled as active by the algorithm.

Although these calculations do provide useful information across the entire tracking period, misclassifications typically only occur at the tails of the rest intervals—that is, most of the uncertainty in estimating rest timing occurs at the beginning and end of the rest interval. We posited that windows of time at the start and end of the rest interval may better capture differences between the three rest interval algorithms relative to hand-scoring. Therefore, we also computed a ‘windowed’ or truncated version of these statistics, where only beginning and ending windows of these intervals are used in the calculation. We considered window sizes up to 240 minutes (four hours) or half of a typical eight-hour rest period. For example, in the calculation of PPV and TPR for a 30-minute window, we filtered the first and last 30 minutes of a rest interval as determined by hand scoring and then calculated PPV and TPR based on only the epochs within those 30-minute intervals.

## RESULTS

### Sample Characteristics

Participant demographic, clinical, and sleep characteristics are described in **Table 2**. The High-Risk and Low-Risk groups had similar demographic features, but the High-Risk group had more psychiatric and sleep pathology. Out of about 14 days of sleep monitoring, on average, participants had >11 valid days of actigraphy tracking, marker presses were completed for rest intervals for 5-7 days, and sleep diaries were available for 10-11 days. A total of 11 out of 51 participants had at least one actigraphy tracking day flagged by actiSleep for review. Less than 1 day of tracking was flagged for review in actiSleep, on average, across participants.

**Table 2.** Demographic, clinical, and sleep characteristics in the full sample and by group (N=51)

| Variable | Full Sample<br>(N=51) |  | High-Risk<br>(n=22) |  | Low-Risk<br>(n=29) |  |
| --- | --- | --- | --- | --- | --- | --- |
|  | Mean<br>or N | SD<br>or % | Mean<br>or N | SD<br>or % | Mean<br>or N | SD<br>or % |
| **Demographics** |  |  |  |  |  |  |
| Age (Yr) | 16.03 | 1.37 | 15.87 | 1.33 | 16.15 | 1.41 |
| Biological Sex (% Female) | 29 | 56.86 | 15 | 68.18 | 14 | 48.28 |
| Ethnicity (% Non-Hispanic) | 47 | 92.16 | 21 | 95.45 | 26 | 89.66 |
| Race |  |  |  |  |  |  |
| White (%) | 36 | 70.59 | 15 | 68.18 | 21 | 72.41 |
| Asian (%) | 0 | 0 | 0 | 0 | 0 | 0 |
| Black (%) | 11 | 21.57 | 5 | 22.73 | 6 | 20.69 |
| Multiple (%) | 4 | 7.84 | 2 | 9.09 | 2 | 6.9 |
| School Grade | 10.16 | 1.29 | 9.82 | 1.33 | 10.41 | 1.21 |
| **Psychiatric Variables** |  |  |  |  |  |  |
| Depression Severity (KDRS) | 2.75 | 4.38 | 4.41 | 5.91 | 1.48 | 2.06 |
| Mania Severity (KMRS) | 1.27 | 2.29 | 2.09 | 3.1 | 0.66 | 1.11 |
| Current Psychiatric Diagnoses |  |  |  |  |  |  |
| Depressive Disorder | 3 | 5.88 | 3 | 13.64 | 0 | 0 |
| Anxiety Disorder | 8 | 15.69 | 8 | 36.36 | 0 | 0 |
| Post-Traumatic Stress Disorder | 1 | 1.96 | 1 | 4.55 | 0 | 0 |
| ADHD | 5 | 9.8 | 5 | 22.73 | 0 | 0 |
| Behavior Disorder | 1 | 1.96 | 1 | 4.55 | 0 | 0 |
| Lifetime Psychiatric Diagnoses |  |  |  |  |  |  |
| Depressive Disorder | 8 | 15.69 | 8 | 36.36 | 0 | 0 |
| Anxiety Disorder | 9 | 17.65 | 9 | 40.91 | 0 | 0 |
| Post-Traumatic Stress Disorder | 1 | 1.96 | 1 | 4.55 | 0 | 0 |
| ADHD | 6 | 11.76 | 6 | 27.27 | 0 | 0 |
| Behavior Disorder | 1 | 1.96 | 1 | 4.55 | 0 | 0 |
| **Sleep Variables** |  |  |  |  |  |  |
| Sleep Quality (PSQI-Past Week) | 3.53 | 2.55 | 4.3 | 3.08 | 2.96 | 1.95 |
| Total Actigraphy Days | 14.12 | 3.91 | 13.64 | 3.97 | 14.48 | 3.89 |
| Valid Actigraphy Days | 11.73 | 4.14 | 12.09 | 4.15 | 11.45 | 4.18 |
| Valid Actigraphy Days with Sleep<br>Diary Rest Onset and Offset Markers | 11 | 5.1 | 10.73 | 5.78 | 11.21 | 4.61 |
| Valid Actigraphy Days with Button-press Rest<br>Onset and Offset Markers | 6.43 | 7.01 | 7.68 | 7.23 | 5.48 | 6.81 |
| Valid Actigraphy days flagged for review | 0.61 | 1.31 | 0.77 | 1.45 | 0.48 | 1.21 |
| Current Sleep Diagnoses |  |  |  |  |  |  |
| Insomnia | 7 | 13.73 | 5 | 22.73 | 2 | 6.9 |
| Hypersomnia | 3 | 5.88 | 2 | 9.09 | 1 | 3.45 |
| Delayed Sleep Phase Disorder | 1 | 1.96 | 0 | 0 | 1 | 3.45 |
| Irregular Sleep-Wake Disorder | 2 | 3.92 | 2 | 9.09 | 0 | 0 |
| Lifetime Sleep Diagnoses |  |  |  |  |  |  |
| Insomnia | 8 | 15.69 | 6 | 27.27 | 2 | 6.9 |
| Hypersomnia | 3 | 5.88 | 2 | 9.09 | 1 | 3.45 |
| Delayed Sleep Phase Disorder | 1 | 1.96 | 0 | 0 | 1 | 3.45 |
| Irregular Sleep-Wake Disorder | 2 | 3.92 | 2 | 9.09 | 0 | 0 |

**Note.** Depressive Disorder = Persistent Depressive Disorder, Major Depressive Disorder, or Unspecified or Other Specified Depressive Disorder; Anxiety Disorder = Generalized Anxiety Disorder, Obsessive Compulsive Disorder, Panic Disorder, Social Phobia, Specific Phobia, Agoraphobia, Separation Anxiety Disorder; PTSD = Post Traumatic Stress disorder; ADHD = Attention Deficit Hyperactivity Disorder; Behavior Disorder = Oppositional Defiant Disorder, Conduct Disorder

### Algorithm Agreement with Hand-Scored Determinations

**Table 3** compares rest interval and sleep measures across each scoring algorithm. Bland-Altman plots of main rest interval onset time (**Fig 3A-B)**, offset time (**Fig 4A-B**), and duration (**Fig 5A-B**) for each algorithm (actiSleep, Activity-Merged, Activity-Only) relative to Hand-Scoring are plotted across the tracking interval. The Bland-Altman plots point to strong mean agreement between each of the three algorithms (actiSleep, Activity-Only, Activity-Merged) with hand-scoring, and low overall proportional bias. However, actiSleep had observably greater agreement with hand-scoring, with less scatter and narrower margins of agreement. Bland-Altman plots for secondary sleep features (**Figures S1**-**S2**) and actograms depicting algorithm differences **(Figures S3-5)** are included in the Supplement.

**Figure 3.**
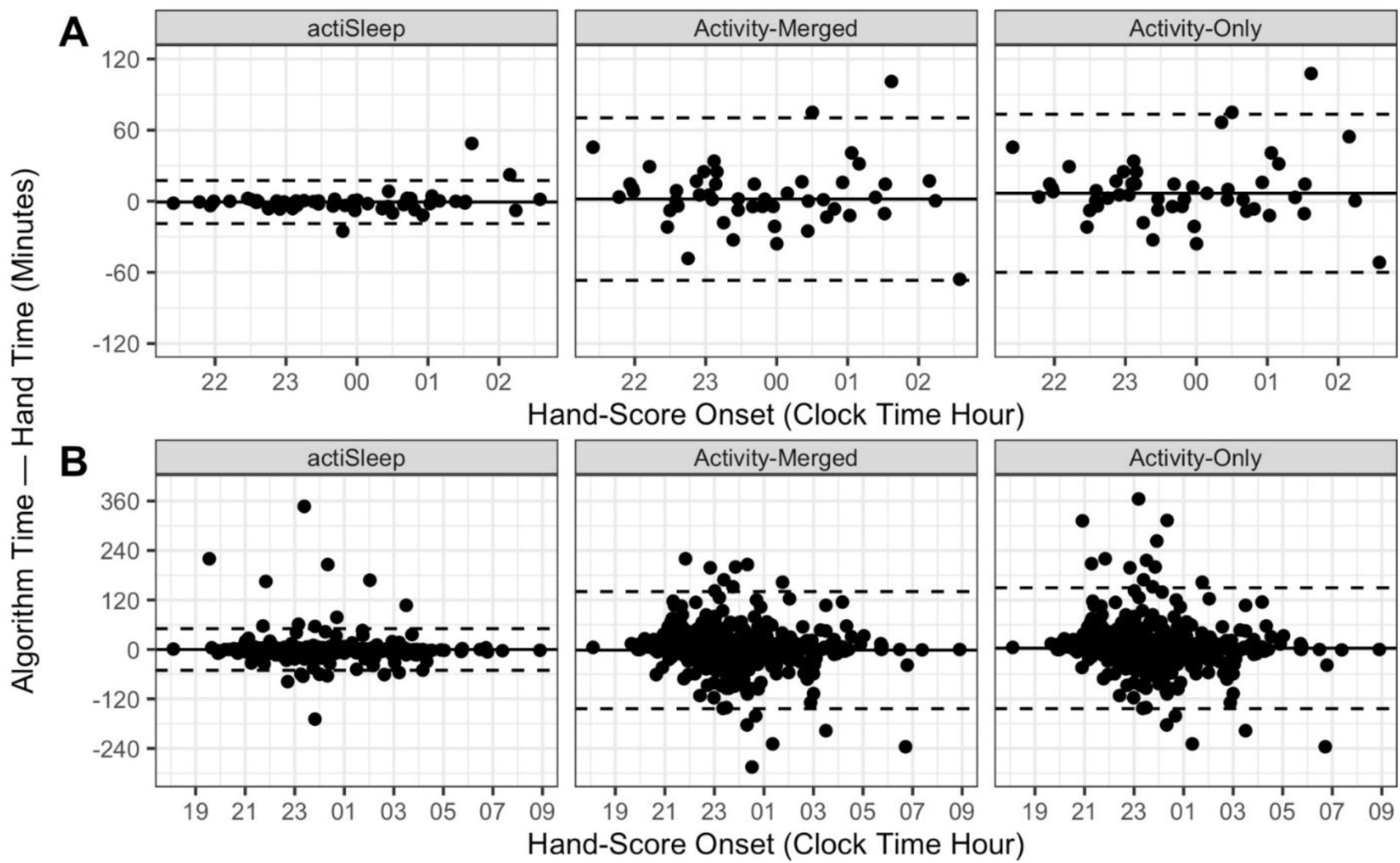
Bland-Altman plots for rest interval onset for each algorithm (actiSleep, Activity-Merged, Activity-Only) relative to Hand-Scoring. The horizontal black line indicates average bias, and the dashed lines reflect the limits of agreement (mean bias ± 1.96 standard deviations). **A.** Rest onset – average across tracking interval; **B.** Rest onset – daily data across the tracking interval

**Figure 4.**
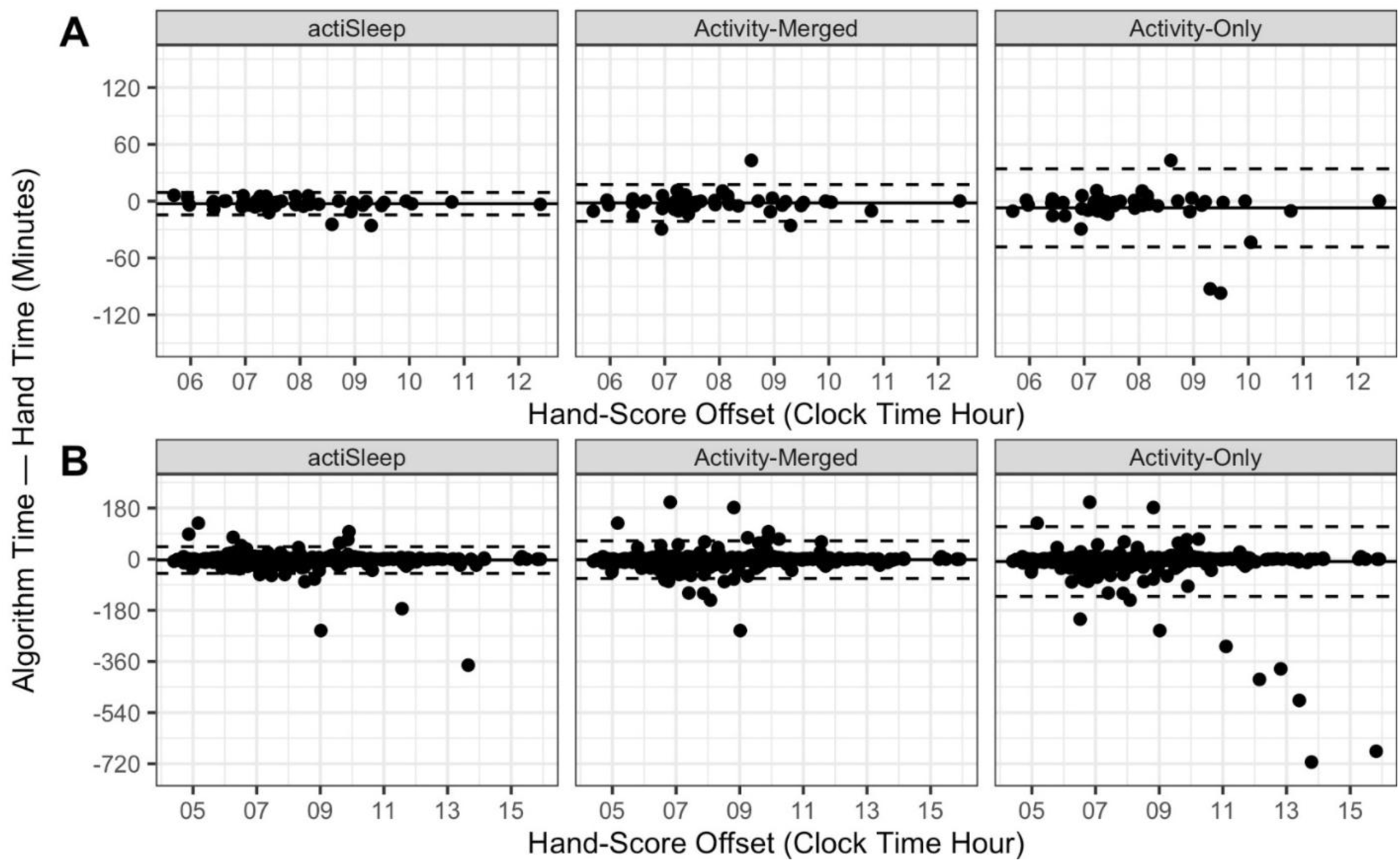
Bland-Altman plots for rest interval offset for each algorithm (actiSleep, Activity-Merged, Activity-Only) relative to Hand-Scoring. The horizontal black line indicates average bias, and the dashed lines reflect the limits of agreement (mean bias ± 1.96 standard deviations). **A.** Rest offset – average across tracking interval; **B.** Rest offset – daily data across the tracking interval

**Figure 5.**
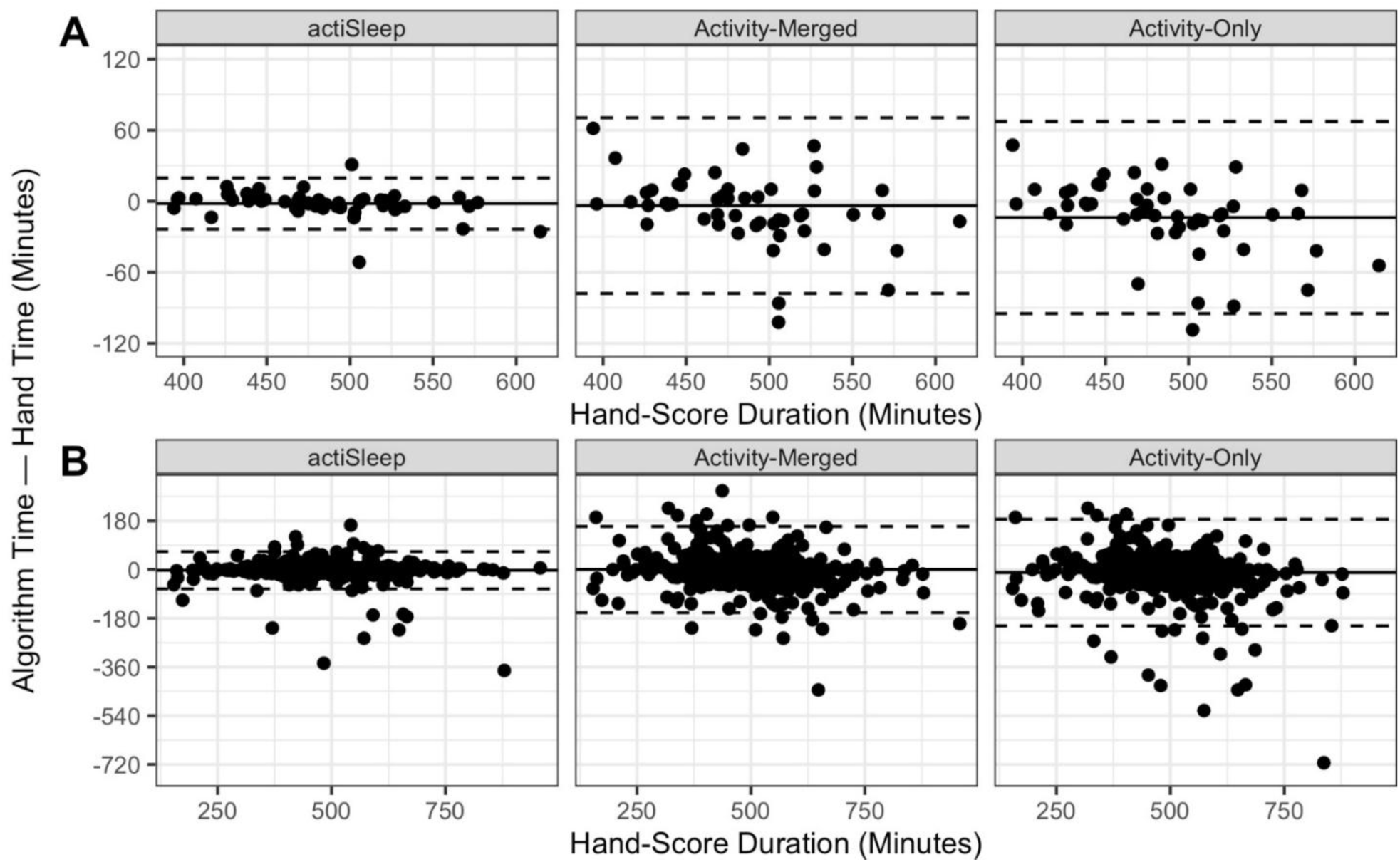
Bland-Altman plots for rest interval duration for each algorithm (actiSleep, Activity-Merged, Activity-Only) relative to Hand-Scoring. The horizontal black line indicates average bias, and the dashed lines reflect the limits of agreement (mean bias ± 1.96 standard deviations). **A.** Rest onset – average across tracking interval; **B.** Rest onset – daily data across the tracking interval

**Table 3.** Actigraphy rest interval and sleep outcomes, by algorithm (N=51).

| Variable, Comparison Algorithm | Hand-Score | Comparison Algorithm | Bias (Algorithm - Hand-Score) |  |  |
| --- | --- | --- | --- | --- | --- |
| | Mean (SD) or Median (IQR) | Mean (SD) or Median (IQR) | Mean or Median $\Delta$ (95% CI) | Statistic | p-val |
| Rest Onset (hh:mm) |  |  |  |  |  |
| actiSleep | 23:48 (01:16) | 23:47 (01:18) | -00:01 (-00:03, 00:02) | -0.52 | 0.603 |
| Activity-Merged |  | 23:50 (01:21) | 00:02 (-00:08, 00:12) | 0.37 | 0.710 |
| Activity-Only |  | 23:54 (01:22) | 00:07 (-00:03, 00:16) | 1.4 | 0.166 |
| Rest Offset (hh:mm) |  |  |  |  |  |
| actiSleep | 07:52 (01:17) | 07:50 (01:16) | -00:03 (-00:04, -00:01) | -3.16 | 0.003 |
| Activity-Merged |  | 07:50 (01:18) | -00:02 (-00:05, 00:01) | -1.36 | 0.181 |
| Activity-Only |  | 07:45 (01:16) | -00:07 (-00:13, -00:01) | -2.4 | 0.020<sup>a</sup> |
| Rest Duration (min) |  |  |  |  |  |
| actiSleep | 484.44 (50.14) | 482.46 (47.8) | -1.99 (-5.09, 1.12) | -1.29 | 0.205 |
| Activity-Merged |  | 480.73 (47.16) | -3.71 (-14.36, 6.93) | -0.7 | 0.487 |
| Activity-Only |  | 470.68 (47.68) | -13.76 (-25.41, -2.12) | -2.37 | 0.022<sup>c</sup> |
| Sleep Onset (hh:mm) |  |  |  |  |  |
| actiSleep | 00:10 (01:18) | 00:13 (01:19) | 00:02 (00:01, 00:04) | 2.63 | 0.011 |
| Activity-Merged |  | 24:05 (01:20) | -00:06 (-00:12, 00:01) | -1.77 | 0.082 |
| Activity-Only |  | 00:08 (01:22) | -00:02 (-00:08, 00:05) | -0.6 | 0.551 |
| Sleep Offset (hh:mm) |  |  |  |  |  |
| actiSleep | 07:36 (01:18) | 07:33 (01:17) | -00:03 (-00:04, -00:01) | -3.38 | 0.001 |
| Activity-Merged |  | 07:35 (01:18) | -00:01 (-00:03, 00:02) | -0.45 | 0.656 |
| Activity-Only |  | 07:30 (01:15) | -00:05 (-00:11, 00:00) | -1.87 | 0.067 |
| Total Sleep Time (min) |  |  |  |  |  |
| actiSleep | 414.56 (41.32) | 413.37 (40.22) | -1.18 (-2.84, 0.47) | -1.44 | 0.156 |
| Activity-Merged |  | 420.01 (42.19) | 5.45 (-0.05, 10.94) | 1.99 | 0.052 |
| Activity-Only |  | 412.43 (43.43) | -2.13 (-8.84, 4.57) | -0.64 | 0.526 |
| Sleep Latency (min) |  |  |  |  |  |
| actiSleep | 17.94 (21.75) | 20.8 (24.06) | 2.29 (1, 4.21) | 1043 | <0.001 |
| Activity-Merged |  | 9.86 (13.74) | -6.01 (-10.28, -2.08) | 341 | 0.004 |
| Activity-Only |  | 9.2 (11.92) | -6.94 (-11.15, -2.87) | 310 | 0.001 |
| WASO (min)<sup>b</sup> |  |  |  |  |  |
| actiSleep | 46.79 (22.83) | 45.27 (22.11) | -1.05 (-1.76, -0.45) | 258 | 0.001 |
| Activity-Merged |  | 48.78 (20.73) | -0.05 (-0.88, 1.03) | 558 | 0.954 |
| Activity-Only |  | 45.2 (23.24) | -0.81 (-1.96, 0.23) | 430.5 | 0.107 |
| Snooze time (min)<sup>b</sup> |  |  |  |  |  |
| actiSleep | 15.42 (9.59) | 15 (9.59) | -0.62 (-1.55, 0.61) | 476 | 0.253 |
| Activity-Merged |  | 14.2 (8.35) | -1.17 (-2.54, 0.08) | 424 | 0.061 |
| Activity-Only |  | 14.2 (7.97) | -1.32 (-2.67, -0.18) | 383.5 | 0.023<sup>c</sup> |
| Sleep Efficiency %<sup>b</sup> |  |  |  |  |  |
| actiSleep | 83.49 (7.24) | 82.89 (7.02) | -0.23 (-0.57, 0.11) | 513 | 0.161 |
| Activity-Merged |  | 84.48 (4.54) | 1.53 (0.7, 2.26) | 1031 | 0.001 |
| Activity-Only |  | 84.82 (4.89) | 1.77 (0.96, 2.52) | 1055 | <0.001 |

**Note.** Grey-shaded values reflect significant pair-wise comparison after Benjamini-Hochburg multiple comparison adjustment; <sup>a</sup>Mean values presented, bias reflects the average mean difference, test statistic is paired t-test; <sup>b</sup>Median values presented, the bias reflects the median of the participant-level differences, and test statistic is Wilcoxon signed-rank tests; <sup>c</sup>Not significant after Benjamini-Hochburg multiple comparison adjustment

### Primary Rest Interval Outcomes

The actiSleep Rest Onset Time and Rest Duration estimates did not significantly differ from Hand-Scoring estimates (average differences < 2-minutes). Similarly, the Activity-Merged Rest Onset Time, Rest Offset Time, and Rest Duration estimates did not significantly differ from Hand-Scoring (average differences < 4 minutes). In addition, Activity-Only algorithm Rest Onset Time, Rest Offset Time, and Rest Duration did not significantly differ from Hand-Scoring (average differences < 14-minutes).

ActiSleep had a significantly earlier Rest Offset Time relative to Hand-Scoring, albeit by relatively a small magnitude (3 minutes).

### Secondary Sleep Outcomes

Overall, the mean and median differences between Hand-Scoring sleep parameters and the three algorithms were of small magnitude (<7 minutes) but some significant differences emerged. Relative to Hand-Scoring, actiSleep slightly underestimated WASO (1.05 minute) and Sleep Offset (3 minutes), and slightly overestimated Sleep Offset (2 minutes) and Sleep Latency (< 3 minutes). Activity-merged and Activity-only both significantly underestimated Sleep Latency (< 7 minutes) and overestimated Sleep Efficiency (< 2 percentage points). None of the three algorithms differed from hand-scoring for Total Sleep Time.

### Positive Predictive Value (PPV) and True Positive Rate (TPR)

For the full rest interval, the epoch-level PPV (>97%) and TPR (>96%) for rest versus active were high on average across the three algorithms (**Table 4**). This high level of agreement is expected, given that the same sleep estimation algorithm was used for all three algorithms and hand-scoring; the middle of the rest interval will significantly overlap and inflate PPV and TPR values. We hypothesized that more notable differences would be observed for the start and end of the rest interval. Indeed, for the 60-minutes and 30-minutes at the start/end of the rest interval, epoch-level comparisons showed greater heterogeneity between algorithms (Figure 6; **Table 4**). actiSleep performed best overall at the 60-minutes and 30-minutes at the tails of the rest interval, which are most critical for defining the rest interval. In the first/last 60-minutes of rest, TPR and PPV remained close to 95% for actiSleep but shifted to the low 90s/high 80s for the other two algorithms. In the first/last 30-minutes of rest, TPR and PPV remained over 90% for actiSleep but shifted to the mid-80s/high 70s for the other two algorithms.

**Figure 6.**
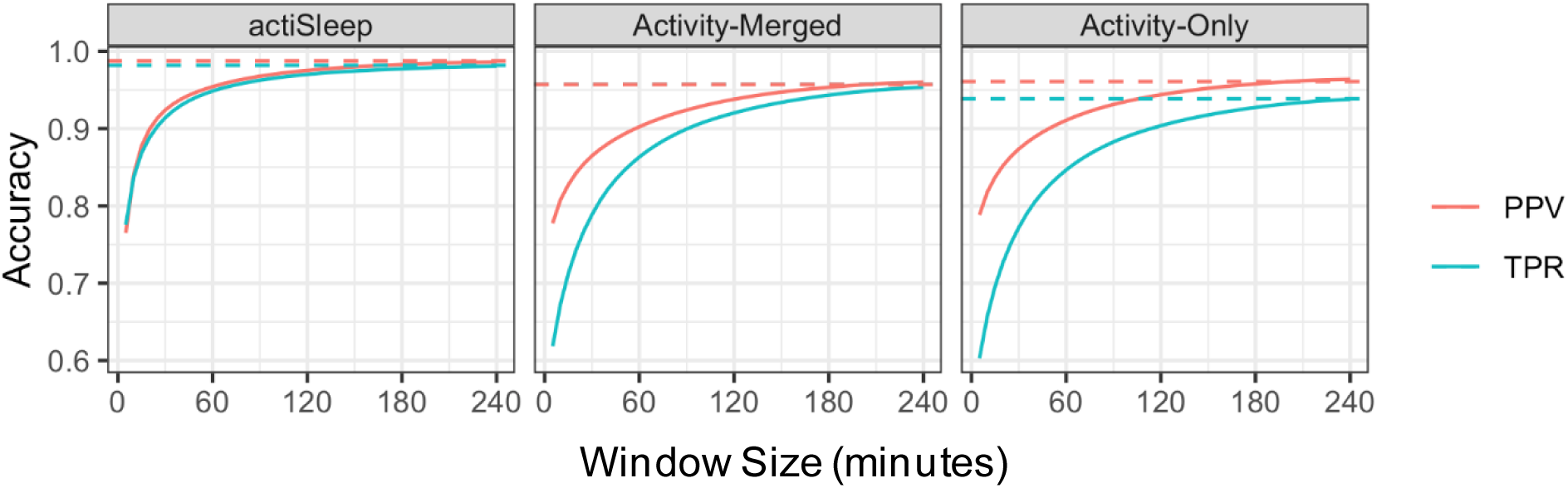
Accuracy as a function of window size in minutes at the start/end of the rest interval. PPV= positive predictive value. TPR = true positive rate for epoch-level sleep detection.

**Table 4.** Positive predictive value and true positive rate of epoch-by-epoch comparisons between hand scoring and three automated rest interval detection algorithms (actiSleep, activity-merged, activity only)

| Algorithm | Positive Predictive Value (PPV) | True Positive Rate (TPR) |
| --- | --- | --- |
|  | % (95% Bootstrap CI) | % (95% Bootstrap CI) |
| **Full rest interval** |  |  |
| actiSleep | 98.75% (98.39%, 99.06%) | 98.19% (97.83%, 98.84%) |
| Activity-Merged | 95.71% (94.93%, 97.62%) | 95.71% (94.51%, 96.67%) |
| Activity-Only | 96.08% (95.31%, 97.84%) | 93.88% (92.33%, 95.57%) |
| **First/Last 60min of Rest** |  |  |
| actiSleep | 95.41% (93.92%, 96.45%) | 94.86% (94.05%, 96.09%) |
| Activity-Merged | 90.24% (88.68%, 93.31%) | 86.34% (82.54%, 88.37%) |
| Activity-Only | 91.08% (89.72%, 93.91%) | 84.66% (80.71%, 86.79%) |
| **First/Last 30min of Rest** |  |  |
| actiSleep | 92.4% (90.28%, 93.81%) | 91.42% (90.36%, 93.08%) |
| Activity-Merged | 86.5% (84.56%, 90.24%) | 78.89% (74.19%, 81.67%) |
| Activity-Only | 87.38% (85.6%, 91.02%) | 77.22% (72.49%, 80.03%) |

### Algorithm Agreement in High-Risk versus Low-Risk Adolescents

**Table S1** and Bland-Altman plots in **Figures S6** describe algorithm comparisons (actiSleep, Activity-Merged, Activity-Only) relative to hand-scoring for High-Risk versus Low-Risk participants. In brief, agreement between all three algorithms with hand scoring was stronger for primary outcomes (rest interval onset, offset, duration) for the Low-Risk group. However, the actiSleep algorithm provided noticeably stronger agreement with less scatter and narrower margins of agreement with hand-scoring in the High-Risk group who had greater sleep and psychiatric disturbances.

## DISCUSSION

Our goal was to develop and perform initial validation of a semi-automated algorithm designed to reproduce hierarchical decision rules applied in manual actigraphy rest interval scoring. To that end, we developed the actiSleep algorithm, which incorporates diary, light, marker presses, and activity in hierarchical estimation of rest interval onset and offset. In a sample of adolescents at high-risk and low-risk for bipolar disorder, rest intervals detected by actiSleep were more consistent with rest intervals estimated by hand-scoring in the overall sample, compared to algorithms relying solely on activity, based on consideration of Bland-Altman plots, PPV, and TPR in aggregate. Improved epoch-level concordance with hand-scoring was most notable at the start and end of the rest intervals for actiSleep. These data provide promising evidence that hierarchical actigraphy scoring rules can be (semi) automated in adolescents, which could greatly improve the scalability of current best-practice approaches for rest interval determination in actigraphy-based sleep estimation.

actiSleep generally improved agreement of Rest Onset, Rest Offset and Rest Duration with hand-scoring relative to activity-based rest interval detection algorithms. Bland-Altman plots of our primary rest interval outcomes clearly depict less between-subject variation in bias across the tracking interval and for daily data. The advantage of actiSleep was most evident for rest onset and duration, which is not surprising given that the highest weighted indicator for rest offset was activity. Even without sleep diary, light, and marker presses, the adapted activity-based algorithm (Activity-Merged) outperformed Activity-Only for our primary rest interval outcomes. Approaches for merging rest intervals have also been proposed in other fully automated activity-based algorithms for rest detection^26^. If activity is the only data stream available for sleep estimation, using an approach akin to our ‘Activity-Merged’ algorithm is a promising middle ground. When examining epoch-level differences in rest versus active classification, actiSleep was notably more consistent with hand-scoring (relative to the activity algorithms) in the ‘tails’ of the rest interval (first and last 30-60 minutes). This outcome is not typically evaluated in validation studies for actigraphy rest interval detection algorithms but is part of the rest interval with greatest ambiguity and the greatest importance for defining rest intervals—which is the cornerstone of all actigraphy-based sleep scoring. Epoch-level accuracy in other studies using the entire rest interval may over-represent the consistency between algorithms.

In exploratory analyses, greater agreement between actiSleep and hand-scoring was particularly evident among the high-risk subgroup of adolescents affected by psychiatric and sleep disorders. In contrast, the performance of the Activity-Merged and Activity-Only algorithms declined in our High-Risk sample for rest interval detection. Accurate actigraphy-based estimation of rest and sleep is historically more challenging in populations with more complex or disrupted sleep patterns^6,27,28^. Our data implies that hierarchical rest interval detection algorithms incorporating multiple data streams may provide the most added value, relative to activity-only algorithms, for establishing rest intervals in populations with disturbed sleep.

There has been a boom in the use of accelerometry in large scale studies. While some large scale cohort studies have deployed standardized hierarchical scoring algorithms^11,12^ these can be highly time-consuming to implement with rigor and, as a result, several large studies use activity-only algorithms that may not perform as well in participants with poor sleep. actiSleep offers a middle-ground to these approaches, bringing principles of hand-scoring to a highly efficient semi-automated algorithm. For the current sample, it took about 20 minutes to implement the rest interval algorithm using hand-scoring for a single 14-day record, corresponding to roughly 17 hours for the entire cohort. However, it only took approximately 1 hour to process the entire cohort in actiSleep (run actiSleep > review and edit flagged days > update rest intervals based on limited manual edits).

We developed the actiSleep hierarchical algorithm based on standardized hand-scoring algorithms^11,13^. Indicator hierarchies, special rules, and flagging may need to be adjusted for specific samples. For example, our approach for flagging ‘long’ sleep intervals for manual review adolescents may have a different threshold for older adults. People may also have different data streams available for use as rest interval indicators, depending on the actigraphy device used. For example, some devices include temperature or heart rate readings. Within actiSleep, users can modify the indicator hierarchy and rest interval definition but should establish concordance with the corresponding hand-scoring method prior to implementing changes to ensure validity. A major advantage of actiSleep is that it has potential to be applied across different actigraphy devices and scoring systems, given the simplicity of the inputs.

The present study should be interpreted considering certain limitations. actiSleep was compared to hand-scoring using a hierarchical algorithm. While hand-scoring actigraphy is currently a best-practice approach, there are inevitably errors when humans implement hierarchical algorithms (e.g., mistakes in the evaluation of indicator hierarchy, ad hoc decisions that do not align with the scoring rules, etc.). Having an imperfect comparator places limits on any measure of agreement. More sophisticated automated machine-learning based algorithms^26,29^ may be other reasonable comparators. In addition, because the actiSleep algorithm was intended to mimic hand-scoring, it does not employ data-driven processes for establishing rest interval indicators (other than a personalized light threshold). Rest interval estimation within actiSleep could be advanced by incorporating machine learning approaches that use all available data streams (e.g., activity, light, marker press, sleep diary) as inputs. While some machine learning and deep learning sleep/rest algorithms exist for actigraphy^5^, they traditionally do not incorporate marker, light, and diary information. This report focused on determination of rest intervals and not sleep/wake estimation by actigraphy per se. While activity-based sleep/wake estimation is not optimal for all samples, tending to over-estimate sleep relative to polysomnography^6–8^, simple heuristic or regression-based algorithms such as HDCZA, Oakley, and Cole-Kripke tend to out-perform more advanced machine learning algorithms for sleep detection^5^. We also did not focus on naps, as there is no standardized algorithm for reliably determining naps, at present. Future developments for sleep/wake detection and rest interval detection (including naps) should ideally progress together. Finally, this initial application of actiSleep used a Philips Spectrum Actiwatch model and sleep estimation in Actiware; since this device is no longer in production, we are working on integrating actiSleep with open-source sleep estimation pipeline GGIR^15^ and implementing it with GeneActiv Original and Ametris (formerly ActiGraph Corporation) devices.

In this initial evaluation of actiSleep performance, we found that actiSleep was a valid semi-automated algorithm relative to hand-scoring among adolescents, performing well in both typically developing adolescents and those affected by sleep and psychiatric pathology. The actiSleep algorithm has potential to improve the efficiency and reliability of rest interval estimation using hierarchical scoring rules, which could be particularly beneficial for large cohort studies. In the future, it will be important to evaluate actiSleep performance in other samples, and in relation to other rest interval detection algorithms. Uptake of standardized automated processing pipelines will be important for improving reproducibility of actigraphy-based sleep studies.

## Data Availability

All data produced in the present study are available upon reasonable request to the authors and with appropriate regulatory approvals.

