## Supplement for "Performance of a Semi-Automated Hierarchical Rest Interval Detection Pipeline (actiSleep) for Wrist Actigraphy in Adolescents"

### Supplemental Results

In addition to the primary Bland-Altman plots on rest and sleep timing, we also considered other sleep characteristics.

In **Figure S1**, we show Bland-Altman plots of sleep latency (**Fig S1A-B)**, sleep efficiency (**Fig S1C-D**), and snooze duration (**Fig S1E-F**) for each algorithm (actiSleep, Activity-Merged, Activity-Only) relative to Hand-Scoring are plotted on average across the tracking interval.

In **Figure S2,** we show Bland-Altman plots of sleep maintenance (**Fig S2A-B)**, total sleep time (**Fig S2C-D**), and wake after sleep onset (**Fig S2E-F**) for each algorithm relative to Hand-Scoring are plotted on averaged across the tracking interval.

In **Figures S3-5**, actograms depict algorithm differences (actiSleep, Activity-Merged, Activity-Only) for 3 participants.

In **Figure S6**, we show Bland-Altman plots of the patient averaged rest onset (**Fig S6A**), rest offset (**Fig S6B**), and rest duration (**Fig S6C**) colored by patient risk level (low versus high) for bipolar disorder.

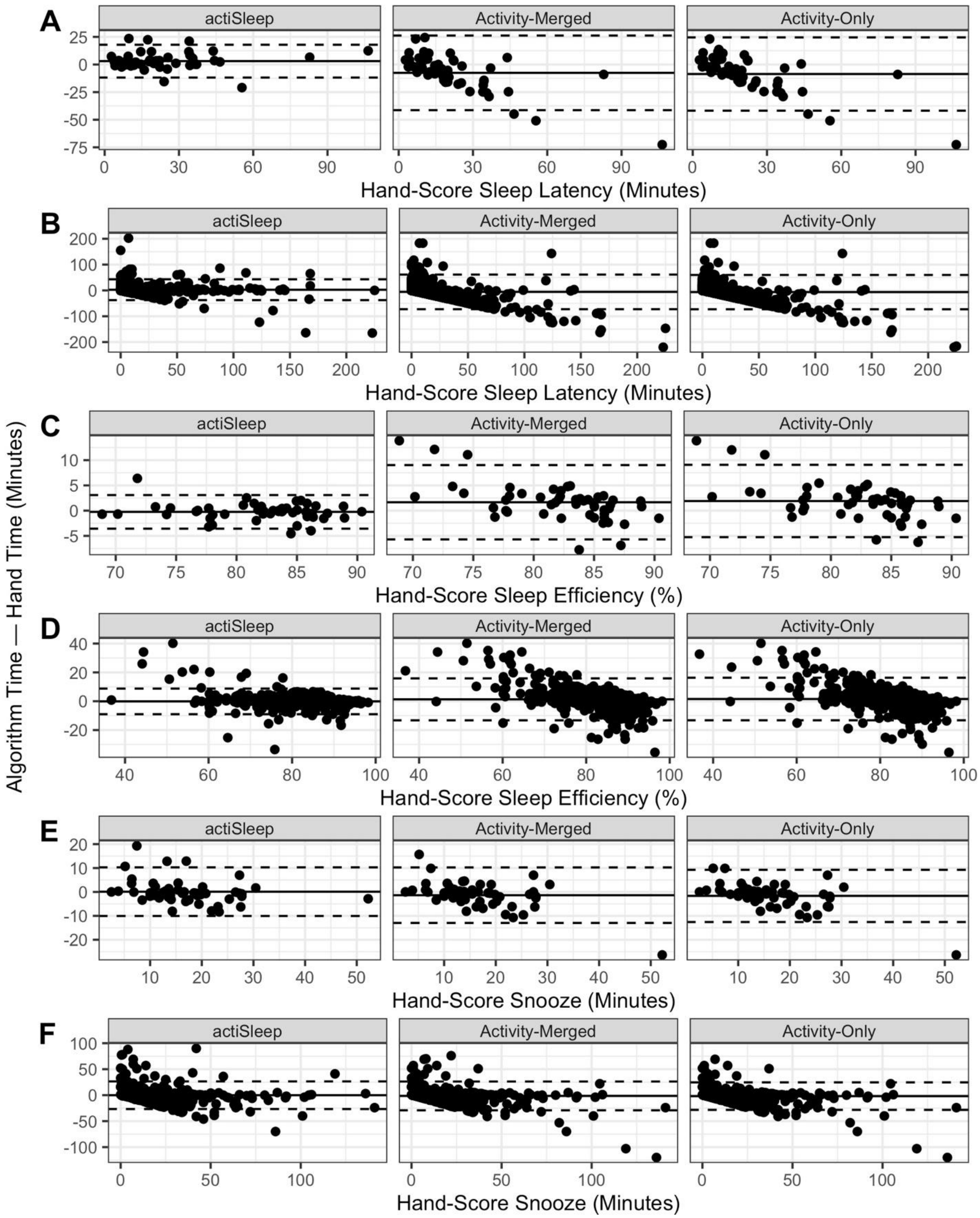


**Figure S1 Caption:** Bland-Altman plots for sleep latency, efficiency, and snooze for each algorithm (actiSleep, Activity-Merged, Activity-Only) relative to Hand-Scoring. **A.** sleep latency average; **B.** sleep latency daily data; **C.** sleep efficiency average; **D.** sleep efficiency daily data; **E.** snooze average; **F.** snooze daily data.

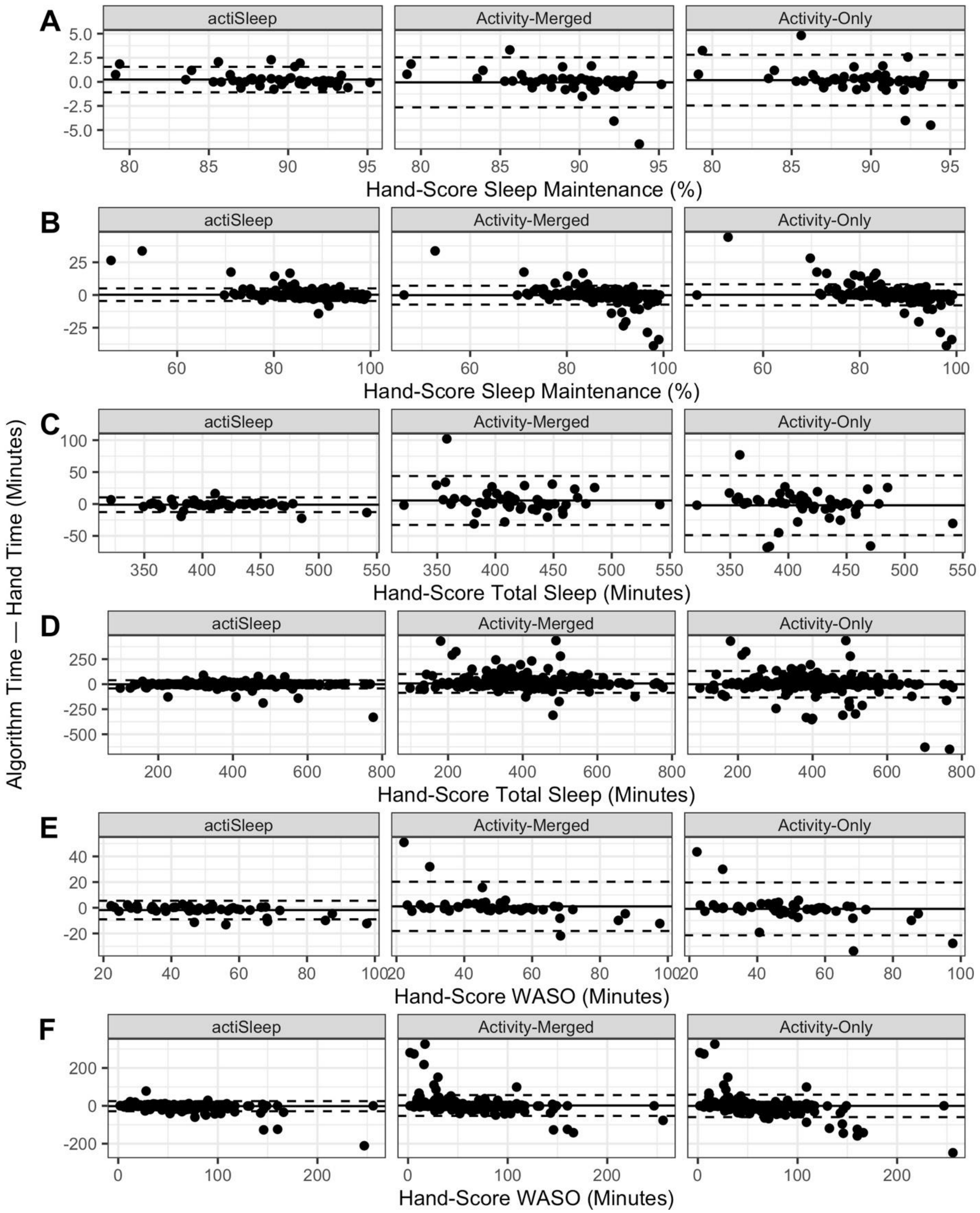


**Figure S2 Caption:** Bland-Altman plots for sleep maintenance, total sleep time, and wake after sleep onset (WASO) for each algorithm (actiSleep, Activity-Merged, Activity-Only) relative to Hand-Scoring. **A.** sleep maintenance average; **B.** sleep maintenance daily data; **C.** total sleep time average; **D.** total sleep time daily data; **E.** WASO average; **F.** WASO daily data.

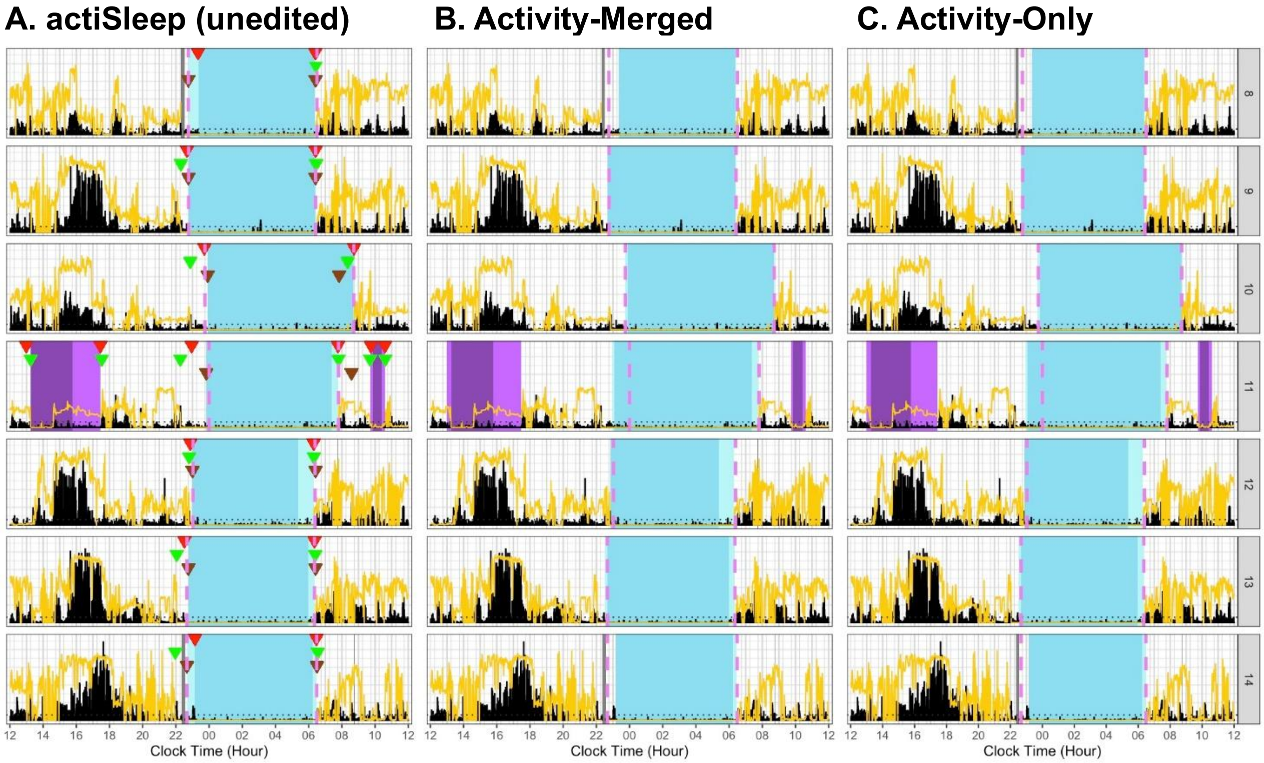


**Figure S3 Caption**. Example actograms generated from actiSleep [**A**] without any flags or excluded days, [**B**] Activity-Merged [**C**], and Activity-Only [**D**] for a single adolescent subject with a stable sleep pattern. There is high agreement between all algorithms and hand scoring. Each row reflects a noon-to-noon tracking day. Activity (black) and light (yellow) are plotted by epoch. Each tracking day, light blue shading reflects a rest interval; the mid-blue shading within the rest interval reflects the sleep interval; purple shading reflects a nap. Rest interval indicators are plotted at the start and end of each rest interval (dark blue = marker press, brown = sleep diary, green = light, red = activity). The pink dashed line indicates hand-scoring rest interval start and end. Navy blue shading reflects a non-wear interval.

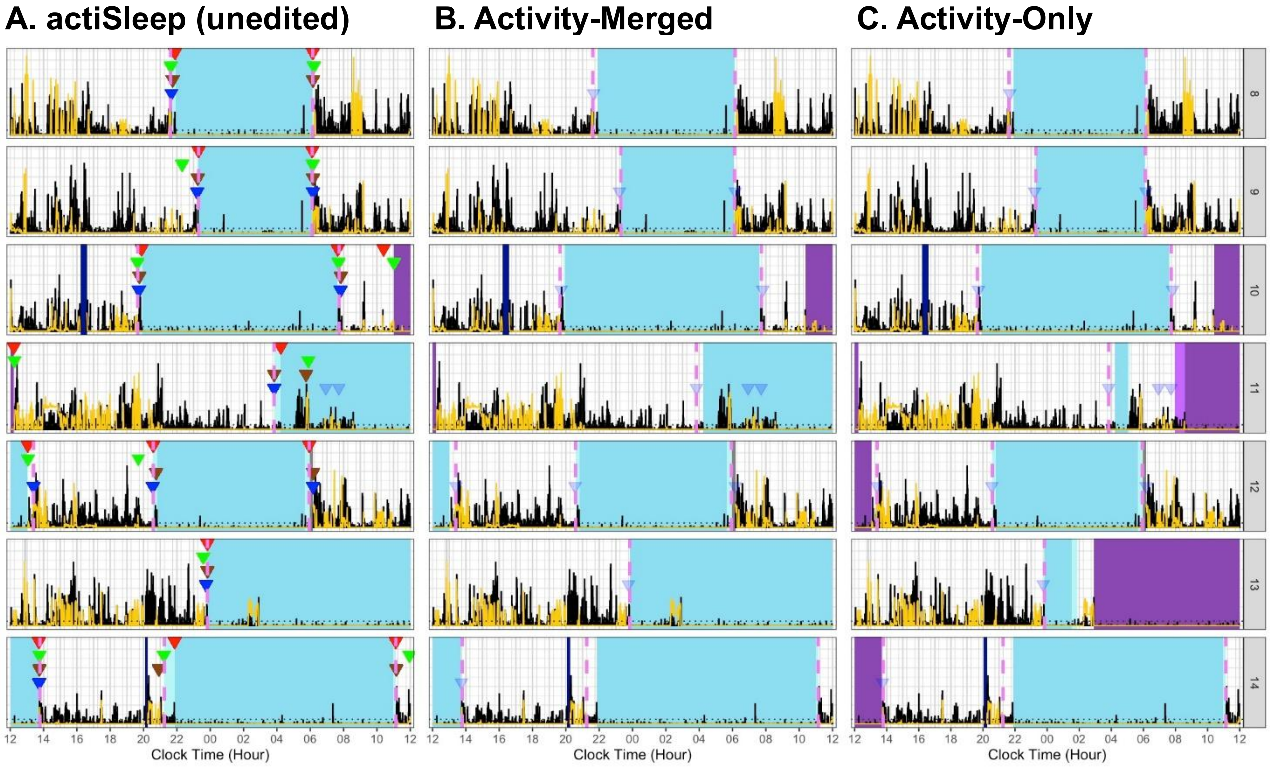


**Figure S4 Caption**. Example actograms generated from actiSleep [**A**] without any flags or excluded days, [**B**] Activity-Merged [**C**], and Activity-Only [**D**] for a single adolescent subject with a variable sleep schedule. There is a high level of agreement between hand-scoring, actiSleep, and Activity-Merged algorithms, with Activity-Only showing differences. Each row reflects a noon-to-noon tracking day. Activity (black) and light (yellow) are plotted by epoch. Each tracking day, light blue shading reflects a rest interval; the mid-blue shading within the rest interval reflects the sleep interval; purple shading reflects a nap. Rest interval indicators are plotted at the start and end of each rest interval (dark blue = marker press, brown = sleep diary, green = light, red = activity). The pink dashed line indicates hand-scoring rest interval start and end. Navy blue shading reflects a non-wear interval.

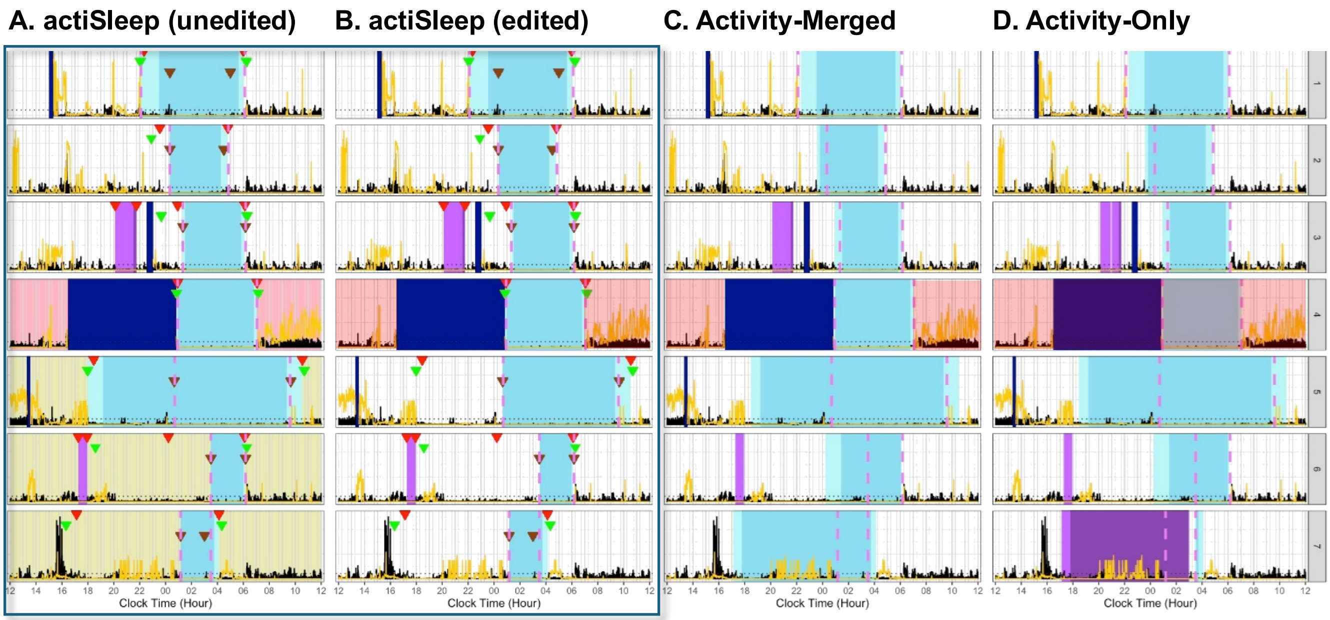


**Figure S5 Caption**. Example actograms generated from actiSleep (before [**A**] and after editing [**B**]), Activity-Merged [**C**], and Activity-Only [**D**] for a single subject. Each row reflects a noon-to-noon tracking day. Light yellow shading of a tracking day reflects a day flagged for manual review. Red shading of a tracking day reflects a day excluded across all algorithms. Activity (black) and light (yellow) are plotted by epoch. Each tracking day, light blue shading reflects a rest interval; the mid-blue shading within the rest interval reflects the sleep interval; purple shading reflects a nap. Rest interval indicators are plotted at the start and end of each rest interval (dark blue = marker press, brown = sleep diary, green = light, red = activity). The pink dashed line indicates hand-scoring rest interval start and end. Navy blue shading reflects a non-wear interval.

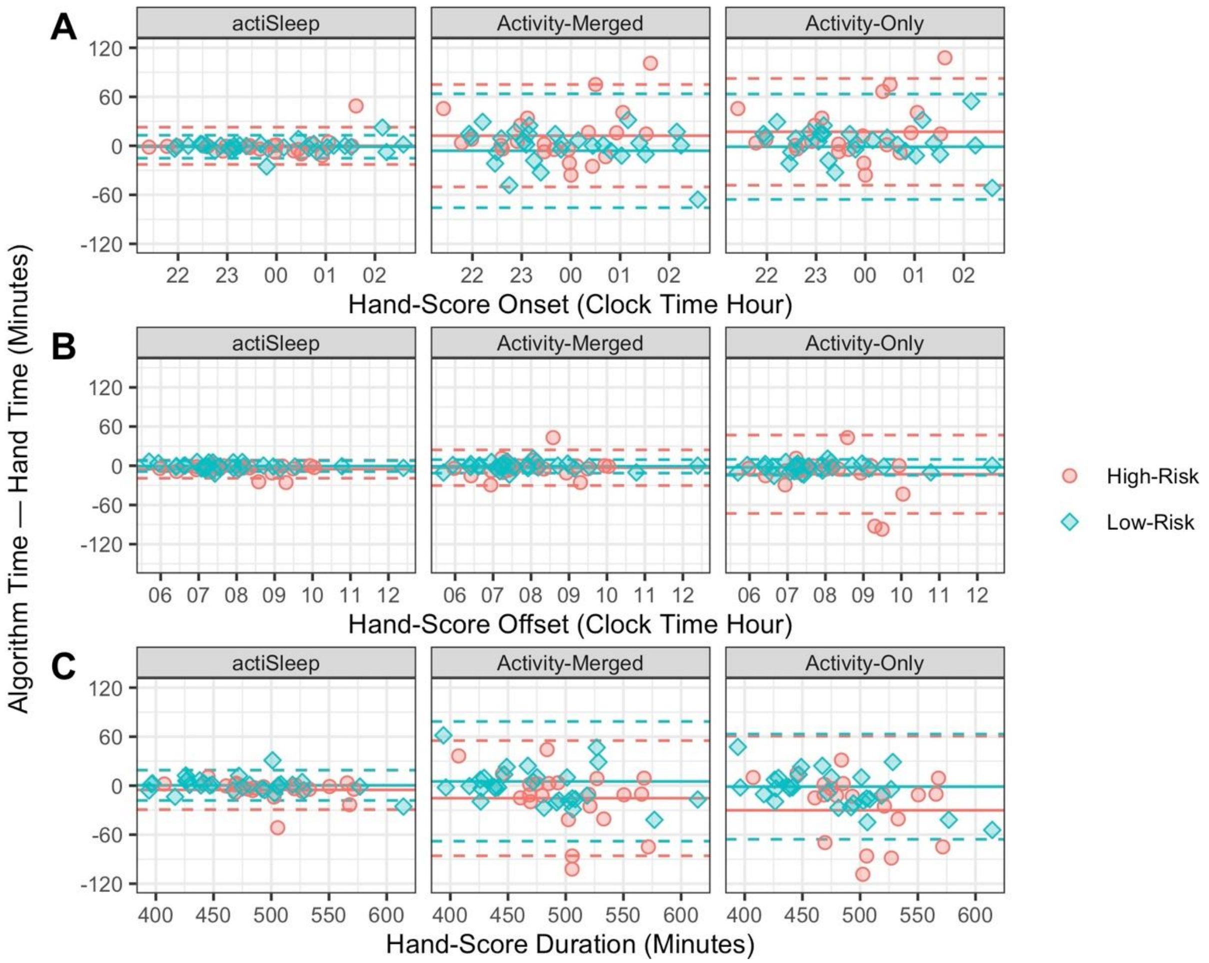


**Figure S6 Caption:** Bland-Altman plots for rest onset, offset and duration for each algorithm (actiSleep, Activity-Merged, Activity-Only) relative to Hand-Scoring colored by risk level. **A.** Rest onset average; **B.** Rest onset average; **C.** Rest offset average.

**Table S1:** Primary actigraphy rest interval outcomes, by risk level and algorithm (N=51).

| *Variable, comparison algorithm* | **High-Risk** | | **Low-Risk** | |
|---|---|---|---|---|
| | *Mean (SD)* | *Mean Δ (95% CI)* | *Mean (SD)* | *Mean Δ (95% CI)* |
| **Rest Onset (hh:mm)** | | | | |
| actiSleep | 23:41 (01:15) | -00:00 (-00:23, 00:23) | 23:52 (01:22) | -00:01 (-00:15, 00:13) |
| Activity-Merged | 23:53 (01:23) | 00:12 (-00:50, 01:15) | 23:47 (01:20) | -00:06 (-01:16, 01:04) |
| Activity-Only | 23:58 (01:26) | 00:17 (-00:48, 01:22) | 23:52 (01:21) | -00:01 (-01:06, 01:03) |
| **Rest Offset (hh:mm)** | | | | |
| actiSleep | 07:56 (01:06) | -00:05 (-00:19, 00:09) | 07:45 (01:23) | -00:01 (-00:09, 00:08) |
| Activity-Merged | 07:58 (01:10) | -00:03 (-00:30, 00:24) | 07:44 (01:24) | -00:01 (-00:11, 00:09) |
| Activity-Only | 07:48 (01:03) | -00:13 (-01:13, 00:47) | 07:43 (01:25) | -00:03 (-00:15, 00:10) |
| **Duration (min)** | | | | |
| actiSleep | 495.2 (40.56) | -5.18 (-29.28, 18.91) | 472.8 (51.2) | 0.44 (-18.16, 19.04) |
| Activity-Merged | 484.98 (43.56) | -15.4 (-85.94, 55.15) | 477.51 (50.24) | 5.15 (-68.16, 78.45) |
| Activity-Only | 470.22 (53.76) | -30.16 (-120.98, 60.65) | 471.03 (43.49) | -1.32 (-65.76, 63.12) |
